# SAMGEP: A Novel Method for Prediction of Phenotype Event Times Using the Electronic Health Record

**DOI:** 10.1101/2021.03.07.21253096

**Authors:** Yuri Ahuja, Chuan Hong, Zongqi Xia, Tianxi Cai

## Abstract

**Objective:** While there exist numerous methods to predict binary phenotypes using electronic health record (EHR) data, few exist for prediction of phenotype event times, or equivalently phenotype state progression. Estimating such quantities could enable more powerful use of EHR data for temporal analyses such as survival and disease progression. We propose Semi-supervised Adaptive Markov Gaussian Embedding Process (SAMGEP), a semi-supervised machine learning algorithm to predict phenotype event times using EHR data.

**Methods:** SAMGEP broadly consists of four steps: (i) assemble time-evolving EHR features predictive of the target phenotype event, (ii) optimize weights for combining raw features and feature embeddings into dense patient-timepoint embeddings, (iii) fit supervised and semi-supervised Markov Gaussian Process models to this embedding progression to predict marginal phenotype probabilities at each timepoint, and (iv) take a weighted average of these supervised and semi-supervised predictions. SAMGEP models latent phenotype states as a binary Markov process, conditional on which patient-timepoint embeddings are assumed to follow a Gaussian Process.

**Results:** SAMGEP achieves significantly improved AUCs and F1 scores relative to common machine learning approaches in both simulations and a real-world task using EHR data to predict multiple sclerosis relapse. It is particularly adept at predicting a patient’s longitudinal phenotype course, which can be used to estimate population-level cumulative probability and count process estimators. Reassuringly, it is robust to a variety of generative model parameters.

**Discussion:** SAMGEP’s event time predictions can be used to estimate accurate phenotype progression curves for use in downstream temporal analyses, such as a survival study for comparative effectiveness research.

## INTRODUCTION

Electronic Health Record (EHR) data collected during the routine delivery of care have in recent years enabled countless opportunities for translational and clinical research.[1–3] Comprising freeform clinical notes, lab results, prescriptions, and various codified features including International Classification of Diseases (ICD) and Current Procedural Terminology (CPT) billing codes, EHR data encode rich information for research. However, a major limitation of EHR data is the lack of precise information on disease phenotypes. Phenotype surrogate features such as ICD diagnosis codes often exhibit dismal specificity that can bias or de-power the downstream study.[4,5] Meanwhile, manual annotation of phenotypes via chart review is laborious and unscalable. These limitations become even more pronounced when the object of interest is the *timing* of clinical events – or equivalently, how clinical status changes over time – which is important for evaluating disease progression. Surrogates of event time derived from EHR codes often exhibit systematic biases, and multiple features may be needed to accurately predict how a phenotype progresses over time.[6-8]

For binary phenotypes, researchers have proposed a variety of unsupervised and semi-supervised methods requiring few-to-no manually-annotated “gold-standard” labels.[9-20] However, very few such methods exist to predict phenotype event times. Chubak et al. developed a rule-based algorithm for breast cancer recurrence that 1) classifies a patient’s overall recurrence status, and 2) for recurrence-positive patients, takes the earliest encounter time of one or more expert-specified codes as the predicted recurrence time.[8] Hassett et al. proposed a similar algorithm averaging the peak times of the pre-specified codes rather than taking the first observed instance.[7] Uno et al. expanded on this by using points of maximal increase in lieu of peak values, and adjusting for systematic temporal biases between the timings of codes and the target phenotype.[6] While these approaches achieve notable performance, they are limited by their 1) reliance on domain expertise to identify predictive codes, 2) inability to utilize more than a handful of codes, and 3) sensitivity to sparsity, a common characteristic of EHR data.

Using more sophisticated machine learning methods to predict phenotype event times can potentially address these limitations. Traditional supervised learning methods such as logistic regression, random forest, and naive Bayes are suboptimal for modeling longitudinal processes as they cannot account for intertemporal associations in both outcomes and features. Recurrent neural networks (RNNs), which are designed for sequence data and well-conditioned to high feature dimensions, have enjoyed particularly widespread application to prediction using EHR data.[21-25] However, these models require large numbers of training labels to achieve stable performance, which is not feasible for phenotypes necessitating manual labeling. Consequently, existing applications of RNNs to EHR-based prediction all use readily available outcome measures such as discharge billing codes, limiting application to phenotypes with reliable codified proxies. In addition, these models are not intuitively interpretable.

On the other end of the spectrum, researchers have developed unsupervised computational models of chronic disease progression that do not require any gold-standard labels. Many of these approaches employ Hidden Markov Models (HMMs) in which latent states represent progressive stages of disease. For instance, Jackson et al. apply a multistage discrete HMM to aneurysm screening, Sukkar et al. apply one to Alzheimer’s disease, and Wang et al. apply a continuous HMM to progression of chronic obstructive pulmonary disease (COPD).[26-29] While these unsupervised models produce promising computational models of disease progression, they may learn latent disease stages that are not clinically relevant.

Using machine learning to predict pre-defined (i.e. not computational) phenotype state progression from EHR data remains a relatively untrodden problem. We draw inspiration from a handful of studies outside the EHR community that use longitudinal physiological measurements to predict sleep state – a binary time series outcome akin to clinical events such as relapse or flare.[30-33] Of particular note, Gao et al. fit a mixed effects logistic regression model to predict sleep state, achieving significantly higher accuracy than naive logistic regression, random forest, or other time-agnostic methods at the expense of requiring a large number of sleep state labels.[30] To efficiently leverage a small number (∼50-100) of labeled and large number of unlabeled longitudinal EHR data to predict the progression of a binary phenotype state, we propose in this paper the Semi-supervised Adaptive Markov Gaussian Embedding Process (SAMGEP) method. Unlike existing event time prediction algorithms, SAMGEP can leverage hundreds of features rather than a handful of surrogates by combining sparse EHR features and their embeddings into dense patient-timepoint embeddings via a novel weighting procedure. It then models the target phenotype state using a Hidden Markov Model with the patient-timepoint embedding progression as a Gaussian Process emission, combining aspects of existing methods to jointly model the evolution of a patient’s phenotype state and EHR feature set over time. SAMGEP enables more powerful application of EHR data to temporal analyses such as survival or disease progression.

## METHODS

The SAMGEP algorithm broadly consists of four steps: (i) assemble time-evolving EHR features predictive of the target phenotype event, (ii) optimize weights for combining raw features and feature embeddings into dense patient-timepoint embeddings, (iii) fit supervised and semi-supervised Markov Gaussian Process (MGP) models to this embedding progression to predict marginal phenotype probabilities at each timepoint, and (iv) take a weighted average of these semi-supervised and supervised predictions with weights determined adaptively to optimize prediction performance. Figure 1 illustrates the overarching SAMGEP procedure.

**Figure 1:**
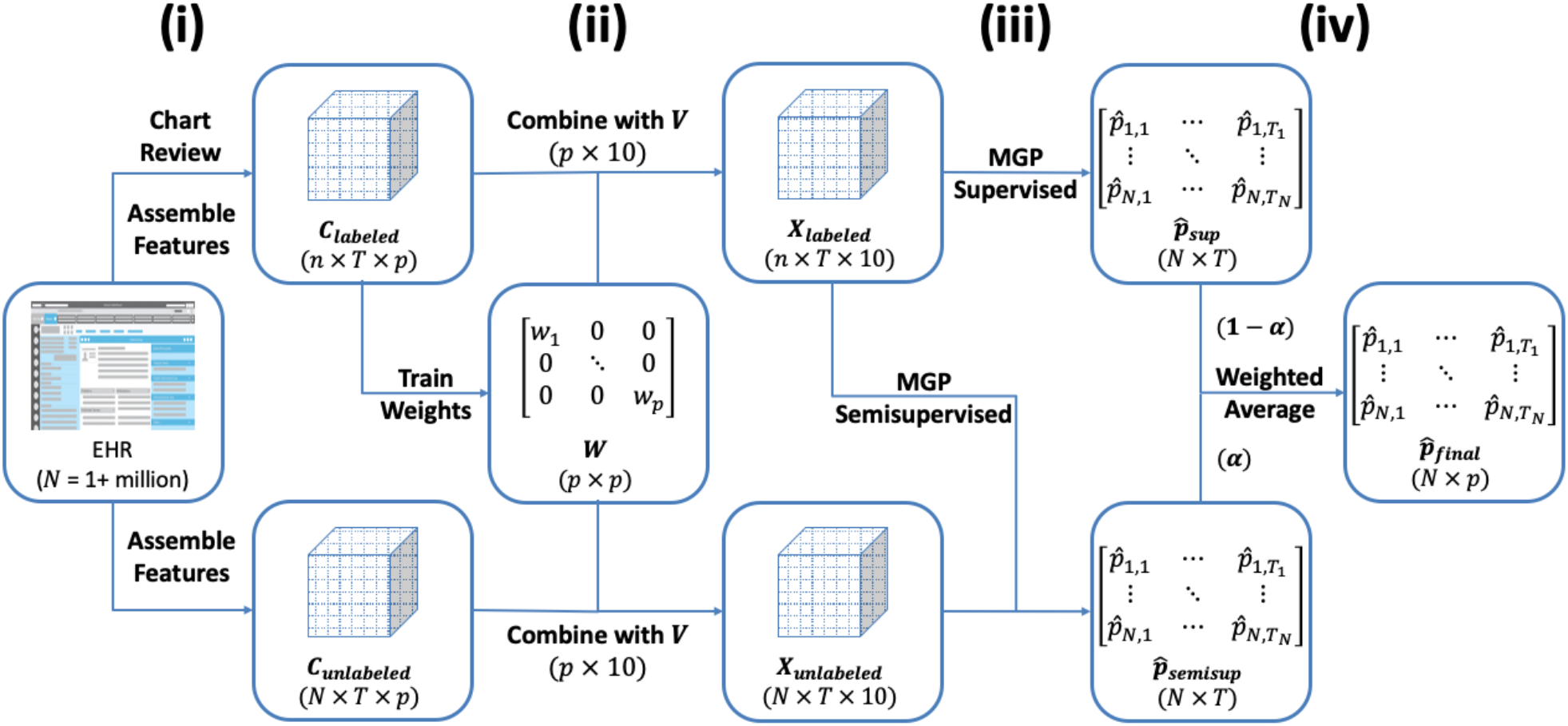
Schematic of the overall SAMGEP algorithm.

### Assembling Predictive Features

Candidate features include log-transformed counts of ICD diagnosis codes or coarser “phenotype” codes (PheCodes), RxNorm drug codes, CPT procedure codes, lab tests, and NLP-curated mentions of relevant concept unique identifiers (CUIs) in a patient’s chart during a given time period. Features can be selected manually or identified automatically via label-free methods such as the surrogate-assisted feature extraction (SAFE) method.[34] In this study, time periods are defined as consecutive non-overlapping 1-month periods starting at the patient’s first PheCode for the target phenotype. Figure 2 depicts the form of raw EHR feature data. Since SAMGEP employs sparse feature weighting to select informative features, it is preferable to aim for inclusivity when assembling features.

**Figure 2:**
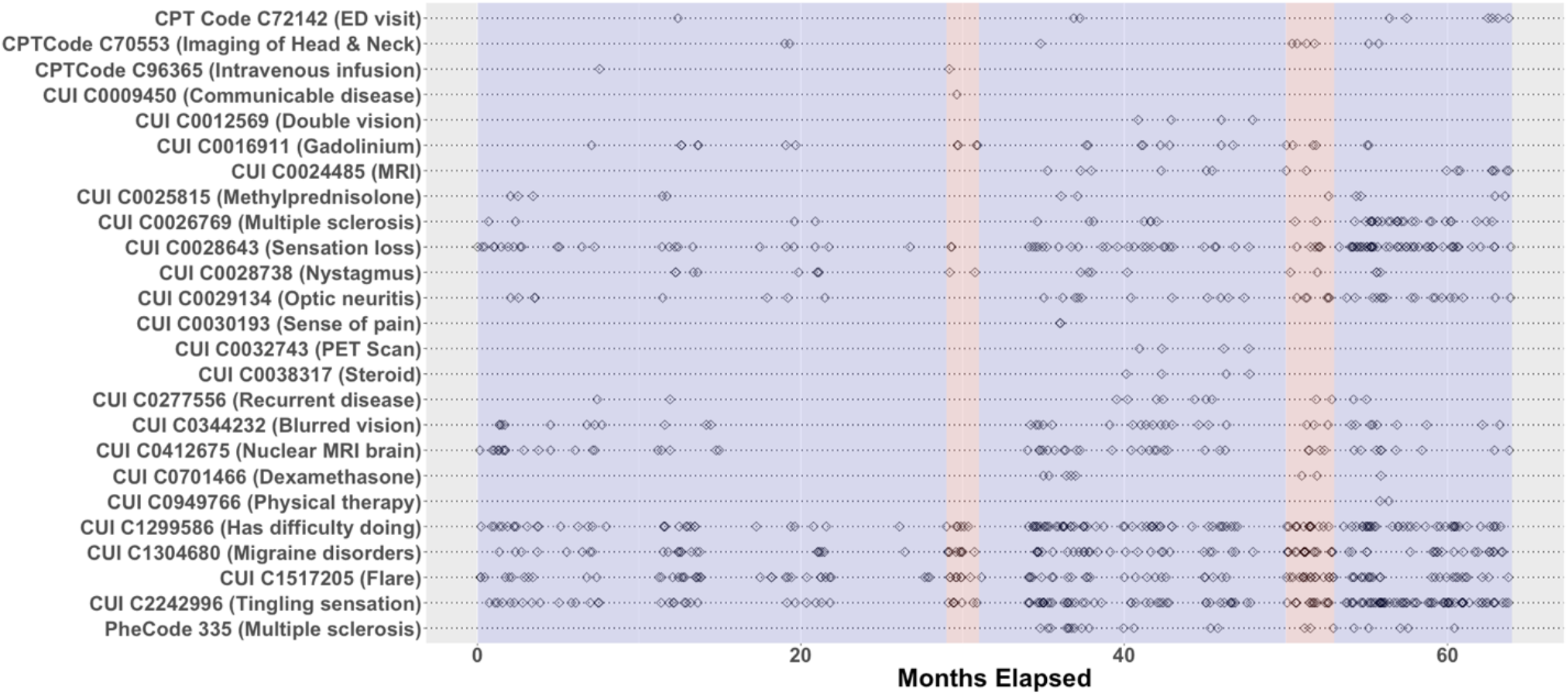
Depiction of the sparsity and temporal irregularity of EHR data. In this study we aim to predict MS relapse event times (red bands) using timestamped EHR feature observations (black diamonds).

Henceforth we let ***V***_*m*×*p*_ denote the matrix of *m*-dimensional embedding vectors of *p* features. We use *i, j*, and *t* to index patients, raw features, and timepoints respectively, and we assume there are a total of *N* patients, and *T*(*i*) timepoints in our dataset. Let ***C***_***i***,***t***_ denote the *p*-dimensional feature vector for patient *i* at timepoint *t*, and *Y*_*i,t*_ ∈ {0,1} denote the phenotype state for patient *i* at timepoint *t*. Moreover, let 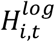 denote patient *i*’s log mean visit count during time period *t* – a measure of healthcare utilization. Finally, we once again assume that we have a limited set of *n* phenotype-labeled patients and a much larger set of *N* − *n* unlabeled ones.

### Producing Patient-Timepoint Embeddings

We compute patient-timepoint embeddings, denoted by ***X***_***i***,***t***_, as a weighted sum over feature embedding vectors:

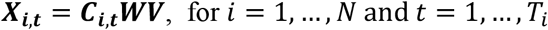

where ***C***_***i***,***t***_ represents the feature log count vector for patient *i* at timepoint *t*, ***W*** is the *p* × *p* diagonal matrix with feature weights on the diagonal, *T*_*i*_ is the total number of timepoints for patient *i, N* is the total sample size, and ***V***_*m*×*p*_ is the matrix of *p m*-dimensional embedding vectors. See the Supplementary Materials for details on how feature embeddings are generated. We choose ***W*** via L1-regularized linear discriminant analysis minimizing:

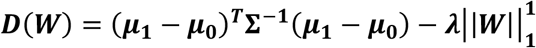

Where

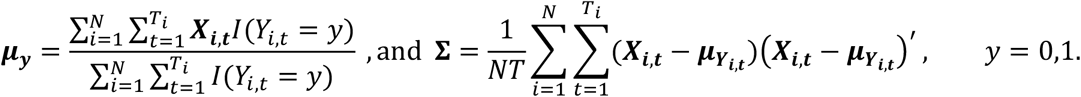

The L1-term imposes sparsity, driving the weights for empirically uninformative features to zero. We optimize ***W*** using projected gradient ascent with the constraint ***W***_1,1_ = **1** to enable identifiability, where without loss of generality we assume that the first feature is a known highly predictive feature. The step-size at each iteration of ascent is chosen by line search. We optimize the regularization hyperparameter *λ* using 10-fold cross-validation within the labeled set.

### Fitting Markov Gaussian Process

Markov Gaussian Process (MGP) is a generative mixture-like model that combines two key assumptions: 1) *Y*_*i*,1_,…, *Y*_*i,T*(*i*)_ follows a discrete time markov process, and 2) the patient embedding vectors over time ***X***_***i***,**1**_,…, ***X***_***i***,***T***(***i***)_ | *Y*_*i*,1_,…, *Y*_*i,T*(*i*)_ follow a Gaussian Process. This generative framework primes SAMGEP for the semi-supervised setting in which we have a large amount of EHR data of which only a limited subset has the longitudinal outcome {*Y*_*i*,1_,…, *Y*_*i,T*(*i*)_} labeled.

#### Discrete Time Markov Process Assumption

We assume a markov process model for *Y*_*i*,1_,…, *Y*_*i,T*(*i*)_ conditional on healthcare utilization *H*_*i*_ such that *P*(*Y*_*i,t*_ = *y*|*Y*_*i*,1_,…, *Y*_*i,t*−1_, *H*_*i*_) = *P*(*Y*_*i,t*_ = *y*|*Y*_*i,t*−1_, *H*_*i*_), where *H*_*i*_ is obtained as the log-count of all encounters throughout a patient’s record. This model can be alternatively specified by two rules:

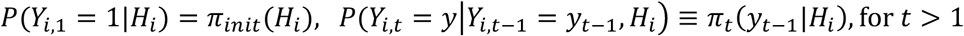

where {*π*_*init*_, *π*_*t*_(*y*_*t*−1_), *t* > 1} are unknown transition probabilities that fully specify the markov model. We further assume that for some ***λ***_markov_ = {*λ*_*init*_, *λ*_0_, *λ*_1_, λ_2_, λ_3_, λ_*H* 0_, λ_*H*_},

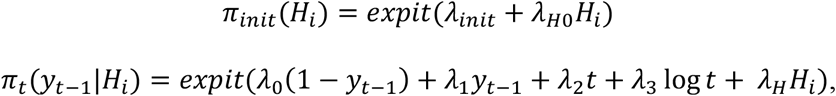

Here we include both linear and log-linear effects for time on *π*_*t*_(*y*_*t*−1_|*H*_*i*_) to better capture how event risk may change over time without overfitting.

#### Gaussian Process Assumption

A Gaussian process is a stochastic process wherein any finite collection of observations follows a multivariate normal distribution. Here, we assume that the patient embeddings over time follow a Gaussian process:

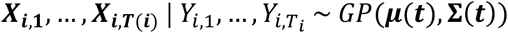

We further specify the mean and covariance functions ***μ***(***t***) and **Σ**(***t***) respectively. For some ***θ***_GP_ = {***μ***_**0**_, ***μ***_**1**_, ***μ***_***H***_, ***μ***_***YH***_, ***μ***_**2**_, ***μ***_**3**_, *σ*_*k*_, *α*_*k*_, *τ*_*k*_, *ρ*_*kl*_, *k* =1,…, *p*; *l* =1,…, *p*}, we assume that:

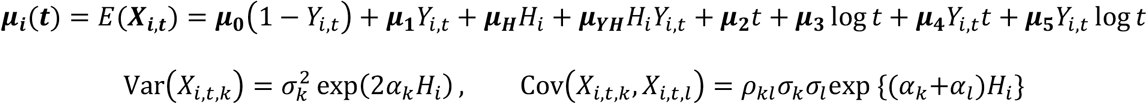

In summary, we assume that patient *i*’s expected embedding at timepoint *t*, ***μ***_***i***_(***t***), is a function of his/her contemporaneous phenotype state *Y*_*it*_, overall healthcare utilization *H*_*i*_, and time *t*. We assume that the marginal variance of embedding component *k* can be represented by some baseline 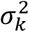 scaled by the effect of healthcare utilization. We denote the correlation between embedding components *k* and *l* as *ρ*_*kl*_, which we assume to be constant over time. Between timepoints, we employ a first-order univariate autoregressive (AR(1)) kernel structure such that the residual at timepoint *t, ϵ*_*i,t,k*_ = *X*_*i,t,k*_ − *E*(*X*_*i,t,k*_|***Y***_***i***_, *H*_*i*_), is a linear function of its temporally previous value *ϵ*_*i,t*−1,*k*_ with autocorrelation coefficient *τ*_*k*_:

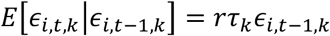

*r* ∈ [0,1] is an autoregression regularization hyperparameter separately trained by 10-fold cross-validation: *r* = 0 ignores intertemporal correlation while *r* = 1 results in undampened autoregression. We chose first-degree autoregression over higher-degree models due to computational ease and mitigation of overfitting.

### Implementation and Inference

MGP is fit via one iteration of an approximating expectation-maximization (EM) algorithm. An EM algorithm iteratively derives an expression for the expected log-likelihood given the current parameter estimates (E-step), and re-computes parameter estimates that maximizes this expected log-likelihood (M-step). Our implementation approximates the expected log-likelihood in the E-step by deriving the marginal posteriors of each latent phenotype state, *Ŷ*_*i,t*_|***X***_***i***_, rather than the more complex joint posterior *Ŷ*_***i***_|***X***_***i***_. Before we fit MGP, we optimize the *r* hyperparameter using 10-fold cross-validation on the labeled set. We then initialize the EM by optimizing the model parameters {***λ***_markov_, ***θ***_GP_} on the labeled set only and using this model to impute labels for the unlabeled set. We refer to predictions made using this initial model as MGP’s supervised estimator 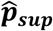. Finally, we re-optimize {***λ***_markov_, ***θ***_GP_} using both observed and imputed phenotype labels. We refer to predictions made using this re-trained model as MGP’s semi-supervised estimator 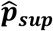. In this study we evaluated the supervised and semi-supervised models on the unlabeled set itself (for which we masked the gold-standard labels), though the models can also be applied to a separate dataset. Specific details of our fitting procedure are supplied in the Supplementary Materials.

### Combining Semi-supervised and Supervised Predictions

Theoretically, semi-supervised generative models such as markov gaussian process should benefit from the additional information in the unlabeled set if the model is correctly specified. However, semi-supervised predictors have been shown to be more sensitive to model misspecification than their supervised counterparts. To mitigate this effect, SAMGEP returns a weighted average of 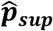 and 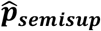, with mixture weight *α* optimized by 10-fold cross-validation maximizing the area under the receiver operating curve (AUC) of held-out *Y*_*i,t*_ predictions:

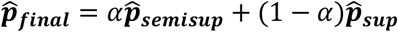

Our results on a real world EHR-based event prediction task demonstrate that this weighted average outperforms both 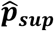 and 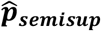 (Figure S1).

### Data and Metrics for Evaluation

#### Simulation Study

To evaluate the SAMGEP algorithm with simulated data, we generated datasets of *p* = 150 count features along with *H* for a variable number of patients, each with a mean of 25 timepoints. To assess our method’s robustness to various model specifications, we varied the following generative parameters: (i) *Y*|*T* where ‘independent’ indicates *Y* ⊥ *T*, ‘correct’ follows SAMGEP’s generative model, and ‘complex’ denotes over-parametrization of *Y*(*T*); and (ii) ***C***|*Y* (marginally lognormal vs. log*t* with 5 df) with intra-temporal correlation matrix fixed at the observed correlation of our real-world dataset and inter-temporal correlation parameter *ρ* varied from 0 to 0.8. We considered *n* = {50,100} and *N* = {1000,5000,20000}. Moreover, we let the number of informative features vary from 5 to 100. Details of our simulation generative mechanisms are supplied in the Supplementary Materials.

#### Real EHR Data Analysis

We collected longitudinal EHR data between January 1, 2006 and December 31, 2016 for 4,706 patients with at least one ICD-9 code for multiple sclerosis (340) from the Partners HealthCare system, which includes Brigham and Women’s Hospital and Massachusetts General Hospital in Boston, MA. We derived neurologist-confirmed multiple sclerosis relapse events and dates for 1,435 patients from the Comprehensive Longitudinal Investigation of Multiple Sclerosis at Brigham and Women’s Hospital (CLIMB) research registry. CLIMB participants have at least one annual clinic visit during the study period. The Partners HealthCare IRB approved the use of both research registry and EHR data.

We defined a relapse event as a clinical and/or radiological relapse. Clinical relapse was defined as having new or recurrent neurological symptoms lasting at least 24 hours without concurrent fever or infection. Radiological relapse was defined as having a new T1-enhancing lesion and/or a new or enlarging T2-FLAIR hyperintense lesion on brain, orbit, or spinal cord MRI.

From the EHR dataset we extracted pertinent demographic information, including age, sex, and race/ethnicity. We also extracted patient-level occurrences of billing codes, including International Classification of Disease 9^th^/10^th^ edition (ICD-9/10) and Current Procedural Terminology (CPT) codes. We mapped ICD codes to PheCodes using the established PheWAS mapping.[35] To mitigate sparsity, we consolidated CPT codes according to groupings defined by the American Medical Association. Finally, from free-text clinical narratives (i.e. discharge summaries) we extracted patient-level occurrences of clinical terms, which we mapped to CUIs using the Natural Language Processing (NLP)-based Narrative Information Linear Extraction (NILE) method.[36]

### Benchmark Methods for Comparison

We considered as benchmarks five supervised methods using the labeled set alone: (i) LASSO-penalized logistic regression [16,17,34,37–39], (ii) random forest (RF) [40,41], (iii) linear discriminant analysis (LDA) [42], and (iv) LSTM-gated recurrent neural network (RNN) [24,39,43,44] trained with raw feature counts ***C***_***i***,***t***_, as well as (v) LDA trained with patient-timepoint embeddings generated without weights 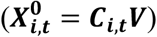, which we refer to as LDA_embed_. In addition, we considered a semi-supervised benchmark: hidden markov model (HMM) [26–29,45,46] with a multivariate gaussian emission trained with the weight-free embeddings 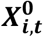. Only HMM and RNN leverage the longitudinal nature of the data, while all other comparator methods train models for predicting *Y*_*t*_ based only on concurrent features (***C***_***i,t***_ or 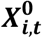) without considering the time sequence. Hyperparameters for LASSO, random forest, and RNN were optimized by 10-fold cross-validation maximizing AUC. As a baseline we also included predictions based only on the multiple sclerosis PheCode (355). While not as meaningful for multiple sclerosis relapse, the closest PheCode is meaningful in situations where the target phenotype is reasonably well coded in the EHR, such as congestive heart failure.

### Evaluation Metrics

To quantify the accuracy of SAMGEP and its comparators’ predictions for the binary phenotype *Y*_*i,t*_, we computed (i) AUC, and (ii) F1 score choosing a cutoff value that achieves 95% specificity. Whereas AUC reflects tradeoff of sensitivity and specificity, F1 score reflects that of sensitivity and positive predictive value. Neither AUC not F1 score consider the sequence of *Ŷ*_*i,t*_ predictions over time for a given patient.

Since the ultimate objective of SAMGEP is to predict the precise timings of phenotype events over the course of a patient’s observed record, as well as the time to first event for survival analysis, we also evaluated the methods’ longitudinal phenotype curve predictions. To this end, we defined the observed all-event counting process (i.e. a patient’s relapse count so far) as *N*_*i*_(*t*) = ∑_*k*≤*t*_ *Y*_*i,k*_(1 − *Y*_*i,k*−1_), where *Y*_*i*,0_ = 0, and the first-event process (i.e. whether a patient has had a relapse yet) as *F*_*i*_(*t*) = 1 − Π_*k*≤*t*_(1 − *Y*_*i,k*_), We defined the all-event counting process this way rather than as *N*_*i*_(*t*) = ∑_*k*≤*t*_ *Y*_*i,k*_ since for an appropriately chosen time window, two consecutive event-positive timepoints {*Y*_*i,k*_ = 1, *Y*_*i,k*+1_ = 1} should correspond to the same event. We evaluated each method’s ability to predict longitudinal phenotype counts by computing the area between *N*_*i*_(*t*) and the predicted counting process 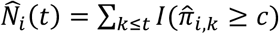, denoted as ABC_count_, where 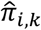 denotes a method’s predicted probability that *Y*_*i,k*_ = 1 and *c* is chosen such that in labeled set

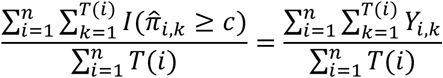

For time to first event, we computed the area between *F*_*i*_(*t*) and the predicted cumulative probability 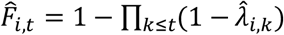, denoted by ABC_cdf_, where λ_*i,k*_ denotes patient *i*’s true hazard at time *k* and 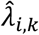 denotes a predictor thereof. Since SAMGEP and HMM jointly model the longitudinal outcome sequence {*Y*_*i*,1_,…, *Y*_*i,T*(*i*)_}, we could use these methods to directly estimate 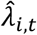. Other methods only predict marginal probabilities 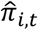 so we assumed that 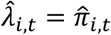, or equivalently that event status labels are independent over time. Rather than report the raw ABC quantities – which don’t have a meaningful scale – we report methods’ percent decrease in the two below those of the null model that sets 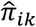 to the prevalence at time *k*:

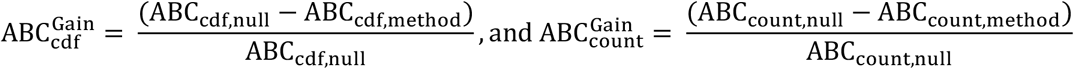

## RESULTS

### Robustness to Data Generative Characteristics

Figure 3 explores SAMGEP and its comparators’ robustness to various generative model specifications. Note that only relative performance between methods, not absolute performance, is meaningful as different generative settings may portend disparate inherent levels of information. Panels A and B reassuringly indicate that SAMGEP, like its comparators, is robust to (A) variations in how phenotype risk changes over time and (B) wide-tailed feature distributions – both realistic misspecifications. Panel C suggests that the benefit of explictly modeling the inter-temporal correlation *ρ* counter-intuitively breaks down at a very high *ρ* of 0.8. At the more realistic *ρ* = 0.4, however, SAMGEP appears more robust than HMM, which assumes that all intertemporal feature correlation is captured by the Markov chain on ***Y***. Panel D suggests that SAMGEP’s use of sparse weights in mapping feature counts ***C*** to patient-timepoint embeddings ***X*** makes the method robust to sparse distribution of information (i.e. 5 or 20 informative features out of 150) at the expense of increased bias in the case of dense information distribution (i.e. 100 informative features), similarly to LASSO. Given that information sparsity is the norm for EHR data, this attribute is well conditioned to EHR modeling. Panel E unsurprisingly demonstrates that larger labeled sets improve predictive AUC, though SAMGEP improves disproportionately between *n* = 50 and *n* = 100. This suggests that while SAMGEP is robust to very low *n*, its true value is unlocked at a labeled set size of ∼100 patients. Finally, panel F shows that SAMGEP singularly benefits from increasing the unlabeled set from *N* = 1000 to *N* = 5000 but not to *N* = 20000. This suggests that SAMGEP is able to effectively extract information from unlabeled patients, though having too many such patients may paradoxically attenuate this benefit.

**Figure 3:**
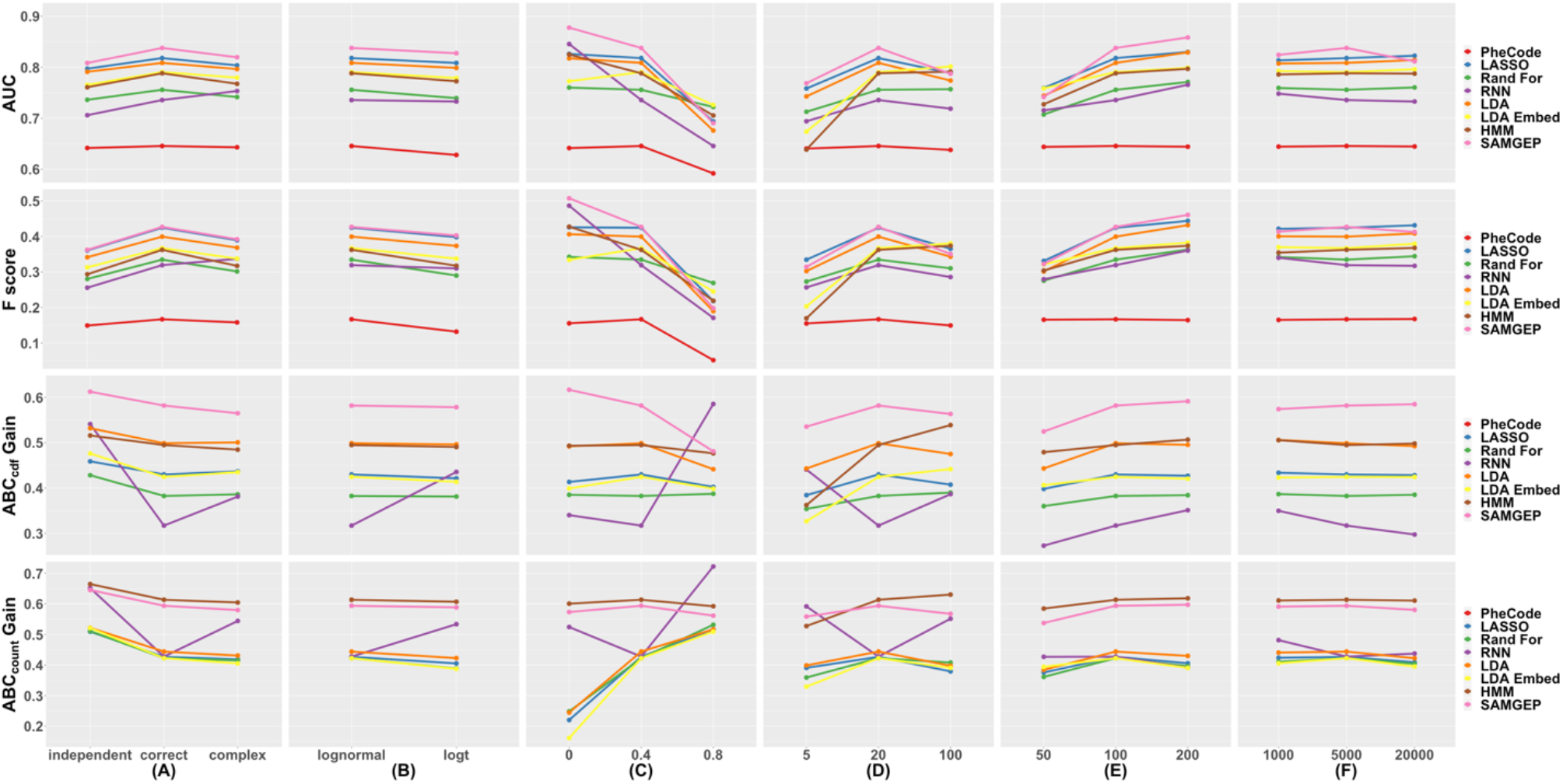
Robustness of SAMGEP and comparator methods’ AUCs, F scores, ABC_cdf_ gains, and ABC_count_ gains to various generative parameters, including the (A) specification of *Y*|*T*, (B) specification of ***X***|*Y*, (C) inter-temporal correlation parameter *ρ*, (D) number of informative (i.e. non-sparse) features, (E) number of labeled patients *n*, and (F) total number of patients *N*. Details of the experiment are delineated in the *Simulation Study* subsection of the Methods.

### Prediction of MS Relapse using Real-World EHR Data

Figure 4 depicts mean AUCs, F1 scores, ABC_cdf_ gains, and ABC_count_ gains for SAMGEP and various comparator methods predicting MS relapse using real-world EHR data. SAMGEP achieved significantly higher mean AUCs and F1 scores than all other methods for both *n* = 50 and *n* = 100 observed labels. RNN on the other hand achieved lackluster AUCs and F1 scores with relatively wide confidence intervals, an unsurprising observation given that such complex neural networks typically require far more than 100 observations to achieve sufficient variance mitigation. SAMGEP also achieved the highest ABC_cdf_ gains, though not significantly so relative to LDA and HMM for *n* = 50, or relative to LDA, HMM, and RNN for *n* = 100. Finally, SAMGEP achieved the highest ABC_count_ gains, though statistically equivalent to RNN and only marginally superior to HMM for both *n* = 50 and *n* = 100. The fact that SAMGEP, HMM, and RNN were among the top performers by both ABC metrics, despite HMM and RNN’s unremarkable AUCs and F1 scores, suggests that jointly modeling {*Y*_*i*,1_,…, *Y*_*i,T*(*i*)_} is singularly beneficial for longitudinal phenotype process prediction. On the other hand, LASSO, random forest, LDA, and LDA_embed_ do not even significantly improve upon the null model counting process estimator, showing that accurately predicting phenotype states at individual timepoints does not necessarily translate into accurate phenotype process prediction.

**Figure 4:**
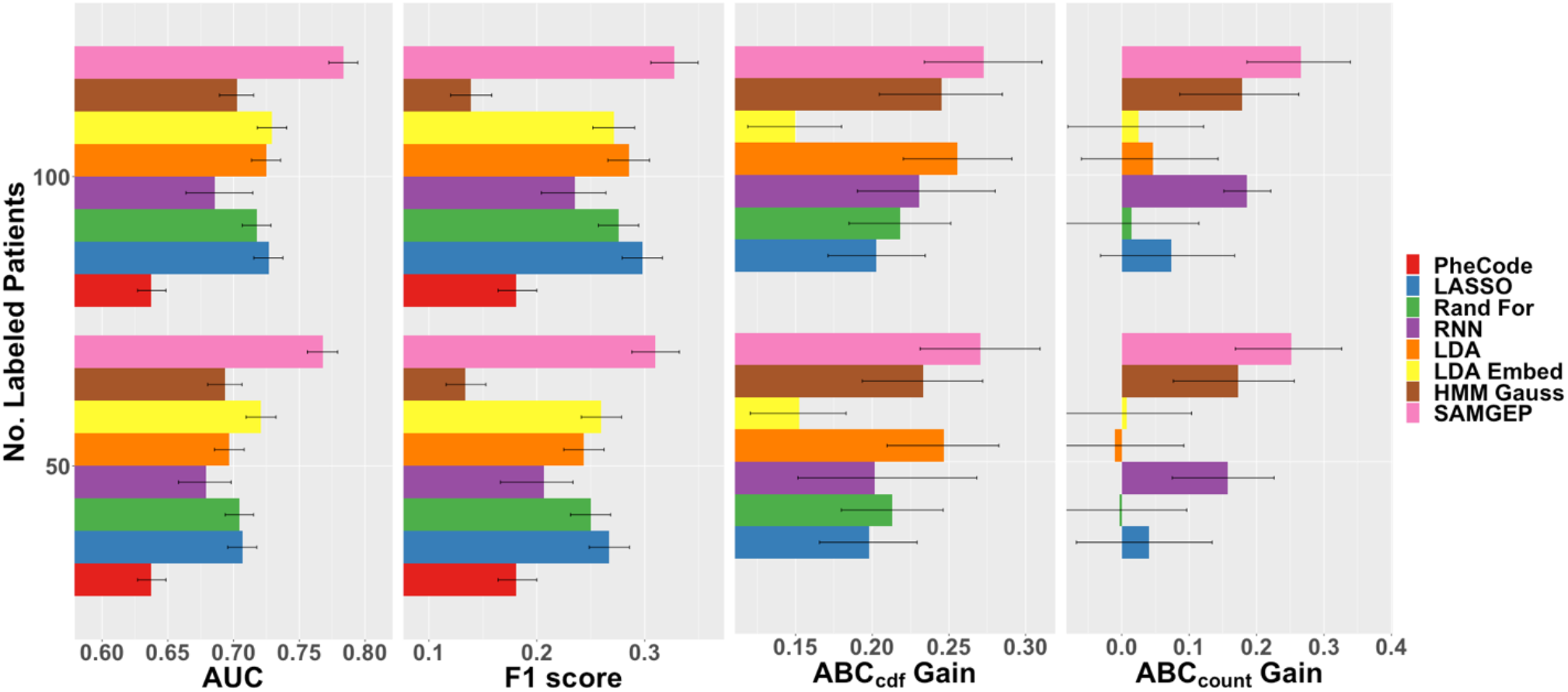
AUCs, F1 scores, ABC_cdf_ gains, and ABC_count_ gains for SAMGEP and various comparator methods predicting MS relapse using real-world EHR data. 95% confidence intervals were empirically estimated by bootstrapping with 100 replicates. See the *Evaluation Metrics* subsection of the Methods for details about the evaluation metrics.

While SAMGEP does not achieve signficant improvement over all comparator methods per all four accuracy metrics, its consistency across metrics is notable. Indeed, while HMM and RNN perform dismally per AUC and F1 score but well per ABC_cdf_ and ABC_count_, and other methods perform better per AUC and F1 score but poorly per ABC_count_, SAMGEP consistently achieves the highest mean accuracy across metrics. It thus demonstrates proficiency at predicting both phenotype states at individual timepoints and phenotype processes across a patient’s record.

Finally, Figure S1 demonstrates that SAMGEP’s mechanism for adaptively weighting MGP’s supervised and semi-supervised predictors improves upon both individual predictors, albeit not significantly. Whereas the supervised predictor tends to achieve higher AUCs and F1 scores, the semi-supervised one tends to perform better per ABC_cdf_ and ABC_count_. Taking a weighted average of the two appears to achieve the best of both worlds.

### Estimation of CDF and Count Process Curves

Figure 5 depicts the estimated population-wide first-relapse CDF, obtained as 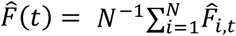, and all-relapse count process, obtained as 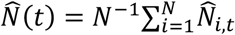, based on the predictions of SAMGEP and comparator methods. Notably, CDF estimation using SAMGEP’s predictions is relatively unbiased, whereas comparator methods all markedly over-estimate the relapse rate. Not shown is the fact that SAMGEP’s confidence intervals are wider than those of LASSO, random forest, LDA, and embedded LDA, explaining why our method does not outperform comparators per the ABC_cdf_ gain metric as significantly as Figure 5 might suggest. The precision of all methods markedly improves from *n* = 50 to *n* = 100, explaining the significant increases we observe in ABC_cdf_ gain despite no apparent improvement in CDF estimation bias.

**Figure 5:**
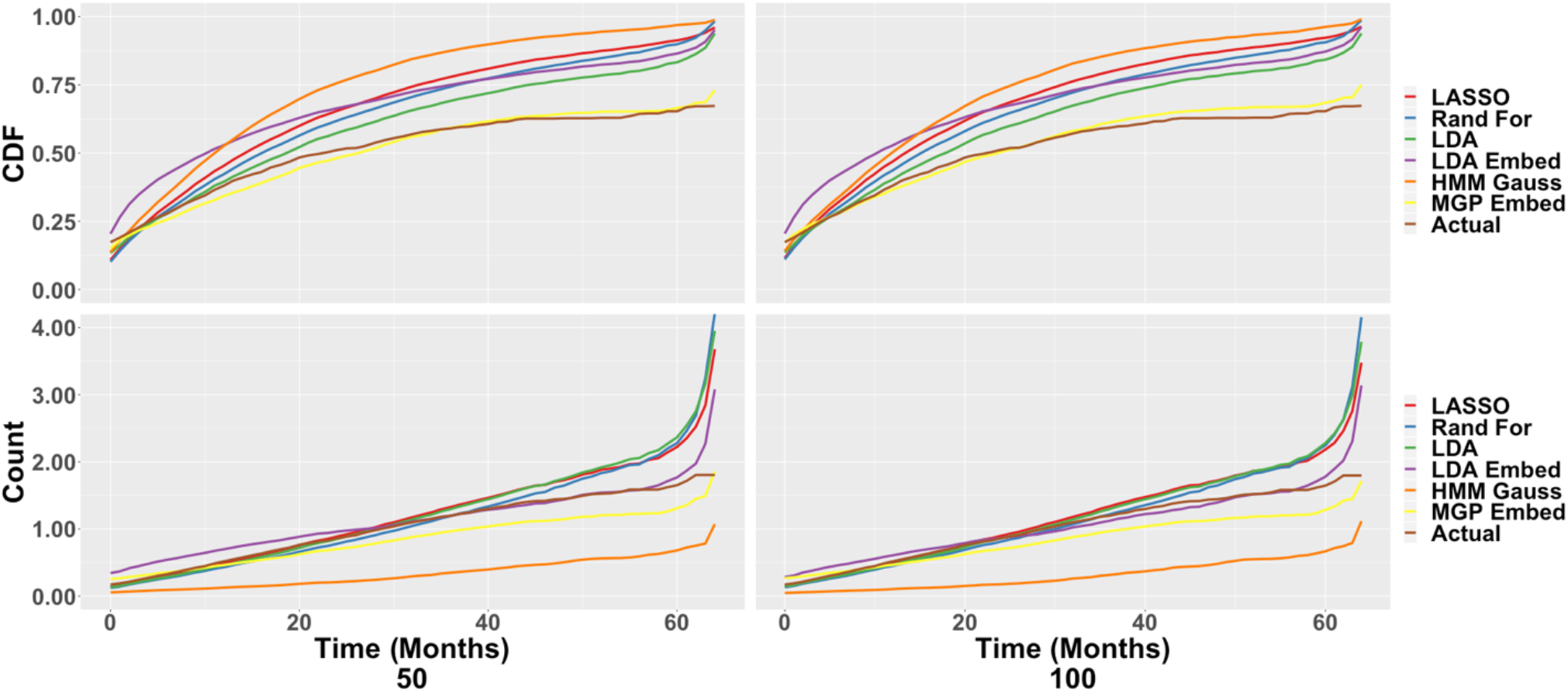
Estimation of population-level MS relapse cumulative probability (top) and count process (bottom) curves as a function of MS disease duration using the predictions of SAMGEP and comparators trained with 50 (left) and 100 (right) labeled patients.

Counting process estimation using SAMGEP’s predictions appears to systematically but slightly underestimate the truth. SAMGEP significantly improves upon comparators later in patients’ disease courses, where all other methods except HMM (which systematically underestimates the true function) exponentially overestimate the mean relapse count. Thus, SAMGEP’s predictions appear to once again improve bias at the expense of increased variance, on the whole significantly improving ABC_count_ gain.

## DISCUSSION

While prediction of binary phenotypes using EHR data is pervasive in the literature, prediction of longitudinal phenotype state progression, or equivalently phenotype event times, remains underdeveloped. As our results demonstrate in concordance with prior studies, accurate prediction of a patient’s overall phenotype status – or even phenotype state at a given timepoint – does not necessarily translate into accurate prediction of phenotype progression. SAMGEP accurately predicts phenotype process functions by effectively leveraging a variety of EHR features with relatively limited expert intervention.

SAMGEP is singularly adept at estimating the cumulative event probability and counting process curves of a binary relapsing-and-remitting phenotype. It thus appears well suited for a study involving survival analysis or estimation of disease progression. For instance, researchers aiming to compare the efficacy of two multiple sclerosis treatments using EHR data could 1) annotate the relapse histories of 50-100 patients via chart review, 2) use SAMGEP to predict relapse probability and count curves for all remaining patients, and 3) use these predictions as features in a survival model such as Cox Proportional Hazards to estimate a population survival function. Further research is warranted to ascertain whether such a workflow affords increased power relative to traditional survival methods using the limited labeled set alone.

The main shortcoming of SAMGEP and other supervised/semi-supervised learning methods is its reliance on gold-standard phenotype event labels. Manually annotating these labels via chart review is particularly labor intensive, requiring an expert to review a patient’s entire chart rather than snapshots thereof. Modifying SAMGEP to handle current status labels – indicators of phenotype status at censor time only – would greatly diminish the manual labor required to utilize SAMGEP for a survival study. Further work is warranted to explore this possibility.

## CONCLUSION

In this study we introduce SAMGEP, a semi-supervised machine learning method that predicts phenotype event times using EHR data and a limited set of gold-standard labels. Singularly adept at estimating event CDF and counting process curves, SAMGEP promises to enable more powerful use of EHR data for epidemological research involving event outcomes, such as survival analysis.

## Data Availability

The datasets used in this paper are proprietary, and as such we unfortunately cannot make them available publicly.

## REFERENCES

[1] I. S. Kohane, S. E. Churchill, and S. N. Murphy, “A translational engine at the national scale: informatics for integrating biology and the bedside,” J. Am. Med. Informatics Assoc., vol. 19, no. 2, pp. 181–185, 2012.

[2] G. Hripcsak and D. J. Albers, “Next-generation phenotyping of electronic health records,” J. Am. Med. Informatics Assoc., vol. 20, no. 1, pp. 117–121, 2012.

[3] R. Miotto, L. Li, B. A. Kidd, and J. T. Dudley, “Deep patient: an unsupervised representation to predict the future of patients from the electronic health records,” Sci. Rep., vol. 6, no. 6, p. 26094 EP, 2016.

[4] K. P. Liao et al., “Electronic medical records for discovery research in rheumatoid arthritis,” Arthritis Care Res, vol. 62, no. 8, pp. 1120–1127, 2010, doi: 10.1002/acr.20184.Electronic.

[5] C. W. Cipparone, M. Withiam-Leitch, K. S. Kimminau, C. H. Fox, R. Singh, and L. Kahn, “Inaccuracy of ICD-9 codes for chronic kidney disease: A study from two practice-based research networks (PBRNs),” J. Am. Board Fam. Med., vol. 28, no. 5, pp. 678–682, 2015, doi: 10.3122/jabfm.2015.05.140136.

[6] H. Uno, D. P. Ritzwoller, A. M. Cronin, N. M. Carroll, M. C. Hornbrook, and M. J. Hassett, “Determining the Time of Cancer Recurrence Using Claims or Electronic Medical Record Data,” JCO Clin. Cancer Informatics, no. 2, pp. 1–10, 2018, doi: 10.1200/cci.17.00163.

[7] M. J. Hassett et al., “Determining the Time of Cancer Recurrence Using Claims or Electronic Medical Record Data,” JCO Clin. Cancer Informatics, vol. 52, no. 10, pp. 1–17, 2015, doi: 10.1097/MLR.0b013e318277eb6f.Validating.

[8] J. Chubak et al., “Administrative data algorithms to identify second breast cancer events following early-stage invasive breast cancer,” J. Natl. Cancer Inst., vol. 104, no. 12, pp. 931–940, 2012, doi: 10.1093/jnci/djs233.

[9] R. J. Carroll et al., “Portability of an algorithm to identify rheumatoid arthritis in electronic health records,” J. Am. Med. Informatics Assoc., vol. 19, no. e1, pp. e162.-e169, 2012.

[10] K. P. Liao et al., “Methods to develop an electronic medical record phenotype algorithm to compare the risk of coronary artery disease across 3 chronic disease cohorts,” PLoS One, vol. 10, no. 8, p. e0136651, 2015.

[11] B. K. Beaulieu-Jones, C. S. Greene, and others, “Semi-supervised learning of the electronic health record for phenotype stratification,” J. Biomed. Inform., vol. 64, pp. 168–178, 2016.

[12] K. M. Newton et al., “Validation of electronic medical record-based phenotyping algorithms: results and lessons learned from the eMERGE network,” J. Am. Med. Informatics Assoc., vol. 20, no. e1, pp. e147.-e154, 2013.

[13] A. N. Ananthakrishnan et al., “Improving case definition of Crohn’s disease and ulcerative colitis in electronic medical records using natural language processing: a novel informatics approach,” Inflamm. Bowel Dis., vol. 19, no. 7, pp. 1411–1420, 2013.

[14] Z. Xia et al., “Modeling disease severity in multiple sclerosis using electronic health records,” PLoS One, vol. 8, no. 11, p. e78927, 2013.

[15] K. P. Liao et al., “Development of phenotype algorithms using electronic medical records and incorporating natural language processing,” bmj, vol. 350, p. h1885, 2015.

[16] J. C. Kirby et al., “PheKB: a catalog and workflow for creating electronic phenotype algorithms for transportability,” J. Am. Med. Informatics Assoc., vol. 23, no. 6, pp. 1046–1052, 2016.

[17] Y. Halpern, Y. Choi, S. Horng, and D. Sontag, “Using anchors to estimate clinical state without labeled data,” in AMIA Annual Symposium Proceedings, 2014, vol. 2014, p. 606.

[18] S. Yu et al., “Enabling phenotypic big data with PheNorm,” J. Am. Med. Informatics Assoc., vol. 25, no. 1, pp. 54–60, 2017.

[19] K. Liao et al., “High-throughput Multimodal Automated Phenotyping (MAP) with Application to PheWAS,” Manuscript, 2019.

[20] Y. Ahuja et al., “sureLDA: A multidisease automated phenotyping method for the electronic health record,” J. Am. Med. Informatics Assoc., Jun. 2020, doi: 10.1093/jamia/ocaa079.

[21] E. Choi, N. Du, R. Chen, L. Song, and J. Sun, “Constructing disease network and temporal progression model via context-sensitive hawkes process,” Proc. - IEEE Int. Conf. Data Mining, ICDM, vol. 2016-Janua, pp. 721–726, 2016, doi: 10.1109/ICDM.2015.144.

[22] D. A. Kaji et al., “An attention based deep learning model of clinical events in the intensive care unit,” PLoS One, vol. 14, no. 2, pp. 1–17, 2019, doi: 10.1371/journal.pone.0211057.

[23] A. Rajkomar et al., “Scalable and accurate deep learning with electronic health records,” npj Digit. Med., vol. 1, no. 1, pp. 1–10, 2018, doi: 10.1038/s41746-018-0029-1.

[24] T. Ruan et al., “Representation learning for clinical time series prediction tasks in electronic health records,” BMC Med. Inform. Decis. Mak., vol. 19, no. Suppl 8, pp. 1–14, 2019, doi: 10.1186/s12911-019-0985-7.

[25] Y. Cheng, F. Wang, P. Zhang, and J. Hu, “Risk prediction with electronic health records: A deep learning approach,” 16th SIAM Int. Conf. Data Min. 2016, SDM 2016, pp. 432–440, 2016, doi: 10.1137/1.9781611974348.49.

[26] C. H. Jackson, L. D. Sharples, S. G. Thompson, S. W. Duffy, and E. Couto, “Multistate Markov models for disease progression with classification error,” Stat., vol. 52, no. 2, pp. 193–209, 2003.

[27] R. Sukkar, E. Katz, Y. Zhang, D. Raunig, and B. T. Wyman, “Disease progression modeling using Hidden Markov Models,” Conf Proc IEEE Eng Med Biol Soc, pp. 2845–2848, 2012.

[28] X. Wang, D. Sontag, and F. Wang, “Unsupervised learning of disease progression models,” Proc. ACM SIGKDD Int. Conf. Knowl. Discov. Data Min., pp. 85–94, 2014, doi: 10.1145/2623330.2623754.

[29] X. Zhou, K. Kang, and X. Song, “Two-part hidden Markov models for semicontinuous longitudinal data with nonignorable missing covariates,” Stat. Med., vol. 39, no. 13, pp. 1801–1816, 2020.

[30] X. Gao, B. Shahbaba, and H. Ombao, “Modeling Binary Time Series Using Gaussian Processes with Application to Predicting Sleep States,” J. Classif., vol. 35, no. 3, pp. 549–579, 2018, doi: 10.1007/s00357-018-9268-8.

[31] E. A. Maharaj, P. D’Urso, and D. U. A. Galagedera, “Wavelet-based Fuzzy Clustering of Time Series,” J. Classif., vol. 27, pp. 231–275, 2010.

[32] B. Zhou, D. E. Moorman, S. Behseta, H. Ombao, and B. Shahbaba, “A Dynamic Bayesian Model for Characterizing Cross-Neuronal Interactions During Decision-Making,” J. Am. Stat. Assoc., vol. 111, no. 514, pp. 459–471, 2014.

[33] F. Wang and A. E. Gelfand, “Modeling Space and Space-Time Directional Data Using Projected Gaussian Processes,” J. Am. Stat. Assoc., vol. 109, no. 508, pp. 1565–1580, 2014.

[34] S. Yu et al., “Surrogate-assisted feature extraction for high-throughput phenotyping,” J. Am. Med. Informatics Assoc., vol. 24, no. e1, pp. e143–e149, 2017, doi: 10.1093/jamia/ocw135.

[35] J. C. Denny et al., “Systematic comparison of phenome-wide association study of electronic medical record data and genome-wide association study data,” Nat. Biotechnol., vol. 31, no. 12, pp. 1102–1111, Nov. 2013, doi: 10.1038/nbt.2749.

[36] S. Yu, T. Cai, and T. Cai, “NILE: Fast Natural Language Processing for Electronic Health Records,” arXiv, pp. 1–23, 2013.

[37] V. M. Castro et al., “Validation of Electronic Health Record Phenotyping of Bipolar Disorder and Controls,” Am. J. Psychiatry, vol. 172, no. 4, pp. 363–372, 2015, doi: 10.1016/j.gde.2016.03.011.

[38] A. E. Anderson, W. T. Kerr, A. Thames, T. Li, J. Xiao, and M. S. Cohen, “Electronic health record phenotyping improves detection and screening of type 2 diabetes in the general United States population: A cross-sectional, unselected, retrospective study,” J. Biomed. Inform., vol. 60, pp. 160–168, 2016, doi: 10.1016/j.physbeh.2017.03.040.

[39] C. Lin et al., “Automatic Prediction of Rheumatoid Arthritis Disease Activity from the Electronic Medical Records,” PLoS One, vol. 8, no. 8, 2013, doi: 10.1371/journal.pone.0069932.

[40] R. Garg, S. Dong, S. Shah, and S. R. Jonnalagadda, “A Bootstrap Machine Learning Approach to Identify Rare Disease Patients from Electronic Health Records Division of Health and Biomedical Informatics, Department of Preventive Medicine, Division of Cardiology, Department of Medicine, Northwestern Unive,” no. 30701070, 2016.

[41] P. L. Teixeira et al., “Evaluating electronic health record data sources and algorithmic approaches to identify hypertensive individuals,” J. Am. Med. Informatics Assoc., vol. 24, no. 1, pp. 162–171, 2017, doi: 10.1093/jamia/ocw071.

[42] S. Yang et al., “Early detection of disease using electronic health records and fisher’s wishart discriminant analysis,” Procedia Comput. Sci., vol. 140, pp. 393–402, 2018, doi: 10.1016/j.procs.2018.10.299.

[43] R. Li et al., “Detection of bleeding events in electronic health record notes using convolutional neural network models enhanced with recurrent neural network autoencoders: Deep learning approach,” J. Med. Internet Res., vol. 21, no. 2, pp. 1–10, 2019, doi: 10.2196/10788.

[44] Z. Yang, M. Dehmer, O. Yli-Harja, and F. Emmert-Streib, “Combining deep learning with token selection for patient phenotyping from electronic health records,” Sci. Rep., vol. 10, no. 1, pp. 1–18, 2020, doi: 10.1038/s41598-020-58178-1.

[45] Z. Sun et al., “A probabilistic disease progression modeling approach and its application to integrated Huntington’s disease observational data,” JAMIA Open, vol. 2, no. 1, pp. 123–130, 2019, doi: 10.1093/jamiaopen/ooy060.

[46] A. Verma, G. Powell, Y. Luo, D. Stephens, and D. L. Buckeridge, “Modeling disease progression in longitudinal EHR data using continuous-time hidden Markov models,” pp. 1–5, 2018.

