## Supplemental Materials for "SAMGEP: A Novel Method for Prediction of Phenotype Event Times Using the Electronic Health Record"

### SUPPLEMENTARY MATERIALS

#### Producing Feature Embeddings

Feature-level embedding vectors were produced by applying singular value decomposition (SVD) to the pointwise mutual information (PMI) matrix of feature occurrence/co-occurrence counts in the Partners HealthCare EHR database. Co-occurrence was defined as the presence of two features in a patient's chart within 1 week of each other. Denoting the marginal occurrence rates of features $i$ and $j$ as $p_i$ and $p_j$ respectively, and the co-occurrence rate of features $i$ and $j$ as $p_{ij}$, the PMI of the two features is defined as

$$pmi(i,j) = \log\left(\frac{p_{ij}}{p_i p_j}\right)$$

Sensibly, PMI is negative if features *i* and *j* are negatively associated, positive if positively associated, and zero if independent. Decomposing the PMI matrix rather than the co-occurrence count matrix is beneficial as PMI normalizes feature prevalence, preventing outsized influence of common features such as the word 'patient' in clinical notes. Since the PMI matrix is symmetric, SVD is simply an eigendecomposition thereof, and the initial embedding vector $\boldsymbol{V_{j,0}}$ for feature $j$ is simply feature $j$'s co-occurrence vector projected onto each PMI eigenvector. For this analysis we took only the first 1,000 eigenvectors, yielding 1,000-dimensional initial embeddings. To further reduce the embedding dimension, we performed PCA on the set of initial embedding vectors corresponding to our assembled features and took the first 10 PC scores for feature *j* as its final embedding vector $\boldsymbol{V_j}$.

#### Fitting MGP using Expectation-Maximization (EM)

*Initialization:* All parameters are initialized on the labeled set. MGP's log-likelihood function can be decomposed into three components: $\log P(\boldsymbol{Y_1})$, $\log P(\boldsymbol{Y_t}|\boldsymbol{Y_{t-1}})$, and $\log f(\boldsymbol{X}|\boldsymbol{Y})$. The first component can be expressed as:

$$\log P(\boldsymbol{Y_1}) = \sum_{i=1}^{N} Y_{i,1} \log \pi_{init} + (1 - Y_{i,t})(1 - \log \pi_{init})$$

where $\pi_{init} = expit(\lambda_{init} + \lambda_{H0} H_i^{log})$. We recognize this is as the objective function of a logistic regression with outcome $\boldsymbol{Y_1}$ and predictor $\boldsymbol{H^{log}}$, and thus fit $\{\lambda_{init}, \lambda_{H0}\}$ using the *glm* package in *R* with binomial outcome and logit link function. Likewise, the second component can be expressed as:

$$\log P(\boldsymbol{Y_t}|\boldsymbol{Y_{t-1}}) = \sum_{i=1}^{N} \sum_{t=2}^{T(i)} Y_{i,t} \log \pi_t + \left(1 - Y_{i,t}\right) \log(1 - \pi_t)$$

where $\pi_t = expit(\lambda_0(1 - Y_{t-1}) + \lambda_1 Y_{t-1} + \lambda_H H_i^{log} + \lambda_t t + \lambda_{logt} \log t)$. This is the objective function of a logistic regression predicting $\boldsymbol{Y_{2:T(i)}}$ from previous phenotype states, $\boldsymbol{H^{log}}$, and $t$, so we fit $\{\lambda_0, \lambda_1, \lambda_H, \lambda_t, \lambda_{logt}\}$ once again using R's *glm* package. Finally, the third component is by design the log-likelihood of a generalized least squares model with outcome $\boldsymbol{X}$ and mean/covariance specified in the "Gaussian Process Assumption" subsection of the Methods section. We fit the mean model and marginal variance parameters $\{\boldsymbol{\mu_0}, \boldsymbol{\mu_1}, \boldsymbol{\mu_H}, \boldsymbol{\mu_{YH}}, \boldsymbol{\mu_t}, \boldsymbol{\mu_{Yt}}, \sigma_{1:m}, \alpha_{1:m}\}$ using the *gls* package in R with first-order autoregression. We compute the maximum likelihood estimator of the intra-temporal correlation parameters $\boldsymbol{\rho} \in R^{m \times m}$ using the $H_i^{\alpha_k}$-normalized residuals of the *gls* fit, $\boldsymbol{\hat{\epsilon}_{i,t}} \in \boldsymbol{R^m}$, as follows:

$$\hat{\rho}_{kl} = \frac{1}{NT\sigma_k \sigma_l} \sum_{i=1}^{N} \sum_{t=1}^{T(i)} \hat{\epsilon}_{i,t,k} \hat{\epsilon}_{i,t,l}$$

Finally, we estimate the inter-temporal autocorrelation parameters $\boldsymbol{\tau} \in R^m$ using component-wise ordinary least squares (OLS) regression of $\boldsymbol{\hat{\epsilon}_{i,t,k}}$ versus $\boldsymbol{\hat{\epsilon}_{i,t-1,k}}$ $\forall\, k \in \{1, \ldots, m\}$. Note that unlike standard vector autoregression, here we assume that $\hat{\epsilon}_{i,t,k} | \hat{\epsilon}_{i,t',k} \perp \hat{\epsilon}_{i,t',l}\ \forall\, t' \neq t, k \neq l$.

*E-step:* Let $\hat{p}_{it} = E[Y_{i,t}|\boldsymbol{X}]$. Again, we estimate the marginal posterior $Y_{i,t}|\boldsymbol{X}$ rather than the joint $\boldsymbol{Y_{i,1:T(i)}}|\boldsymbol{X}$, which dramatically improves computational efficiency at the expense of being unable to re-optimize intertemporal parameters. Since both Markov Process and first-order autoregression assume that a timepoint is independent of the past and future conditional on its neighboring timepoints, we can accurately approximate $\hat{p}_{i,t}$ as $E\left[Y_{i,t}\middle|\boldsymbol{X_{i,t-1}},\boldsymbol{X_{i,t}},\boldsymbol{X_{i,t+1}}\right]$ rather than $E\left[Y_{i,t}\middle|\boldsymbol{X_{i,1:T(i)}}\right]$:

$$\hat{p}_{i,t}=\frac{\sum_{u=0}^{1}\sum_{w=0}^{1}P\left(Y_{i,t-1}=u\right)P\left(Y_{i,t}=1|Y_{i,t-1}=u\right)P\left(Y_{i,t+1}=w|Y_{i,t}=1\right)f(\boldsymbol{X_{i,t-1}},\boldsymbol{X_{i,t}},\boldsymbol{X_{i,t+1}}|Y_{i,t-1},Y_{i,t},Y_{i,t+1})}{\sum_{u=0}^{1}\sum_{v=0}^{1}\sum_{w=0}^{1}P\left(Y_{i,t-1}=u\right)P\left(Y_{i,t}=v|Y_{i,t-1}=u\right)P\left(Y_{i,t+1}=w|Y_{i,t}=v\right)f(\boldsymbol{X_{i,t-1}},\boldsymbol{X_{i,t}},\boldsymbol{X_{i,t+1}}|Y_{i,t-1},Y_{i,t},Y_{i,t+1})}$$

Note that $P\left(Y_{i,t-1}=u\right)$ here is a marginal probability, independent of $\{\boldsymbol{X_i},Y_i\}_{1:t-2}$. While this is misspecified, it is faster and indeed achieves higher test set accuracy in our real-world EHR example than jointly estimating $E\left[\boldsymbol{Y_{i,1:T(i)}}\middle|\boldsymbol{X_{i,1:T(i)}}\right]$. $f\left(\boldsymbol{X_{i,t-1}},\boldsymbol{X_{i,t}},\boldsymbol{X_{i,t+1}}\middle|Y_{i,t-1},Y_{i,t},Y_{i,t+1}\right)$ is simply the density of the multivariate normal specified in the "Gaussian Process Assumption" subsection. Finally, for the endpoints $t=\{1,T(i)\}$, we respectively omit $\left\{\boldsymbol{X_{i,t-1}},Y_{i,t-1}\right\}$ and $\left\{\boldsymbol{X_{i,t+1}},Y_{i,t+1}\right\}$ in the above computation and predict $\hat{p}_{i,t}$ as above using the remaining two timepoints.

*M-step:* Re-optimizing the model parameters follows a similar procedure to initialization. The expected log-likelihood function can be decomposed into three components: $E[\log P(\boldsymbol{Y_1})]$, $E[\log P(\boldsymbol{Y_t}|\boldsymbol{Y_{t-1}})]$, and $E[\log f(\boldsymbol{X}|\boldsymbol{Y})]$. Note that since we only estimate marginal rather than joint posteriors in the E-step, we cannot derive a closed-form expression for the second component and therefore maintain the transition parameters $\{\lambda_0,\lambda_1,\lambda_H,\lambda_t\}$ at their initial values. Likewise, we cannot re-infer the autocorrelation parameters $\tau_{1:m}$ and thus maintain them at their initial values as well. The first component can be expressed as:

$$E[\log P(\boldsymbol{Y_1})]=\sum_{i=1}^{N}\hat{p}_{i1}\log\pi_{init}+(1-\hat{p}_{i1})(1-\log\pi_{init})$$

We recognize this as the objective function of a weighted logistic regression with outcome $[\boldsymbol{0_N},\boldsymbol{1_N}]$, predictor $[\boldsymbol{H^{log}},\boldsymbol{H^{log}}]$, and observation weights $[\boldsymbol{1}-\boldsymbol{\hat{p}_1},\ \boldsymbol{\hat{p}_1}]$, and thus refit $\{\lambda_{init},\lambda_{H0}\}$ accordingly using the *glm* package. Similarly, the third component becomes the log-likelihood of a generalized least squares model with outcome $[\boldsymbol{X},\boldsymbol{X}]$, mean $[\boldsymbol{\mu}|\boldsymbol{Y}=\boldsymbol{0},\ \boldsymbol{\mu}|\boldsymbol{Y}=\boldsymbol{1}]$, covariance specified in the "Gaussian Process Assumption" subsection of the Methods, and observation weights $[\boldsymbol{1}-\boldsymbol{\hat{p}_1},\ \boldsymbol{\hat{p}_1}]$. We can thus refit $\{\boldsymbol{\mu_0},\boldsymbol{\mu_1},\boldsymbol{\mu_H},\boldsymbol{\mu_{YH}},\boldsymbol{\mu_t},\boldsymbol{\mu_{Yt}},\sigma_{1:m},\alpha_{1:m}\}$ using the *gls* package in R with first-order autocorrelation. Finally, we re-estimate $\boldsymbol{\rho}$ using the $H_i^{\alpha_k}$-normalized residuals of the weighted *gls* fit, $[\hat{\boldsymbol{\epsilon}}_{\boldsymbol{i,t,Y0}}, \hat{\boldsymbol{\epsilon}}_{\boldsymbol{i,t,Y1}}] \in \boldsymbol{R^{2m}}$, where $\hat{\boldsymbol{\epsilon}}_{\boldsymbol{i,t,Y0}}$ denotes the residuals for observations where $\boldsymbol{Y = 0}$ and $\hat{\boldsymbol{\epsilon}}_{\boldsymbol{i,t,Y1}}$ where $\boldsymbol{Y = 1}$:

$$\hat{\rho}_{kl} = \frac{1}{NT\sigma_k\sigma_l}\sum_{i=1}^{N}\sum_{t=1}^{T(i)}(1-\hat{p}_{i,t})\hat{\epsilon}_{i,t,Y0,k}\hat{\epsilon}_{i,t,Y0,l} + \hat{p}_{i,t}\hat{\epsilon}_{i,t,Y1,k}\hat{\epsilon}_{i,t,Y1,l}$$

**Simulation Data Generative Mechanisms**

In our simulation study we generated datasets via a four-step procedure: for each patient, generate (i) the total number of timepoints $T_i$ and (ii) the initial phenotype state $Y_{i,0}$; for timepoints $2\!:T_i$ generate (iii) $Y_{i,t}|Y_{i,t-1}$; and (iv) generate longitudinal feature counts $\boldsymbol{C_i}|\boldsymbol{Y_i}$. We vary the following generative parameters:

(1) The mechanism of $Y|T$, where 'independent' indicates that $Y \perp T$ (i.e. $Y_{i,t} \sim Bern(\pi_0\{H_i\}) \; \forall \; i,t)$ , 'correct' follows SAMGEP's generative mode (i.e. $Y_{i,t} \sim Bern(\pi_{i,t}), \pi_{i,t} = expit\{\lambda_0(1-y_{t-1}) + \lambda_1 y_{t-1} + \lambda_2 t + \lambda_3 \log t + \lambda_H H_i\}$), and 'complex' denotes over-parametrization of $Y(T)$ (i.e. $Y_{i,t} \sim Bern(\pi_{i,t}), \pi_{i,t} = expit\{\lambda_0(1-Y_{t-1}) + \lambda_1 y_{t-1} + \lambda_2 t + \lambda_3 \log t + \lambda_H H_i + \lambda_{02}(1-Y_{t-1})t + \lambda_{12} y_{t-1} t + \lambda_{03}(1-Y_{t-1})\log t + \lambda_{13} Y_{t-1} \log t + \lambda_{0H}(1-Y_{t-1})H_i + \lambda_{1H} Y_{t-1} H_i + \lambda_{2H} H_i t + \lambda_{3H} H_i \log t\}$), with generative parameters $\boldsymbol{\lambda}$ optimized using our real-world MS relapse dataset;

(2) The marginal distribution of $\boldsymbol{C_i}|\boldsymbol{Y_i}$, where 'lognormal' indicates that marginally $\log C_{i,j,t}\, |Y_{i,t} \sim N(\alpha_{0,j}(1-Y_{i,t}) + \alpha_{1,j}Y_{i,t}, \sigma^2)$ and 'log-t' that $\log C_{i,j,t}\, |Y_{i,t} \sim t(\alpha_0(1-Y_{i,t}) + \alpha_{1,j}Y_{i,t}, 5df)$;

(3) The inter-temporal correlation parameter $\rho$ of $\boldsymbol{C}|\boldsymbol{Y}$, where $cor(C_{i,t}, C_{i,s}|\boldsymbol{Y}) = \rho^{|t-s|}$

(4) The number of observed phenotype labels $n$;

(5) The total number of patients $N$;

(6) The number of informative features, where the generative $\beta$ coefficients of any non-informative features are set to 0.

In summary, we generated datasets per the mechanisms outlined in the following table:

| $\boldsymbol{Y \mid T}$ | $\boldsymbol{C \mid Y}$ | $\boldsymbol{\rho}$ | $\boldsymbol{n}$ | $\boldsymbol{N}$ | $\boldsymbol{n}$**Informative** |
|---|---|---|---|---|---|
| Correct | Lognormal | 0.4 | 100 | 5000 | 20 |
| Complex | Lognormal | 0.4 | 100 | 5000 | 20 |
| Independent | Lognormal | 0.4 | 100 | 5000 | 20 |
| Correct | Log-t | 0.4 | 100 | 5000 | 20 |
| Correct | Lognormal | 0 | 100 | 5000 | 20 |
| Correct | Lognormal | 0.8 | 100 | 5000 | 20 |
| Correct | Lognormal | 0.4 | 50 | 5000 | 20 |
| Correct | Lognormal | 0.4 | 200 | 5000 | 20 |
| Correct | Lognormal | 0.4 | 100 | 1000 | 20 |
| Correct | Lognormal | 0.4 | 100 | 20000 | 20 |
| Correct | Lognormal | 0.4 | 100 | 5000 | 5 |
| Correct | Lognormal | 0.4 | 100 | 5000 | 100 |

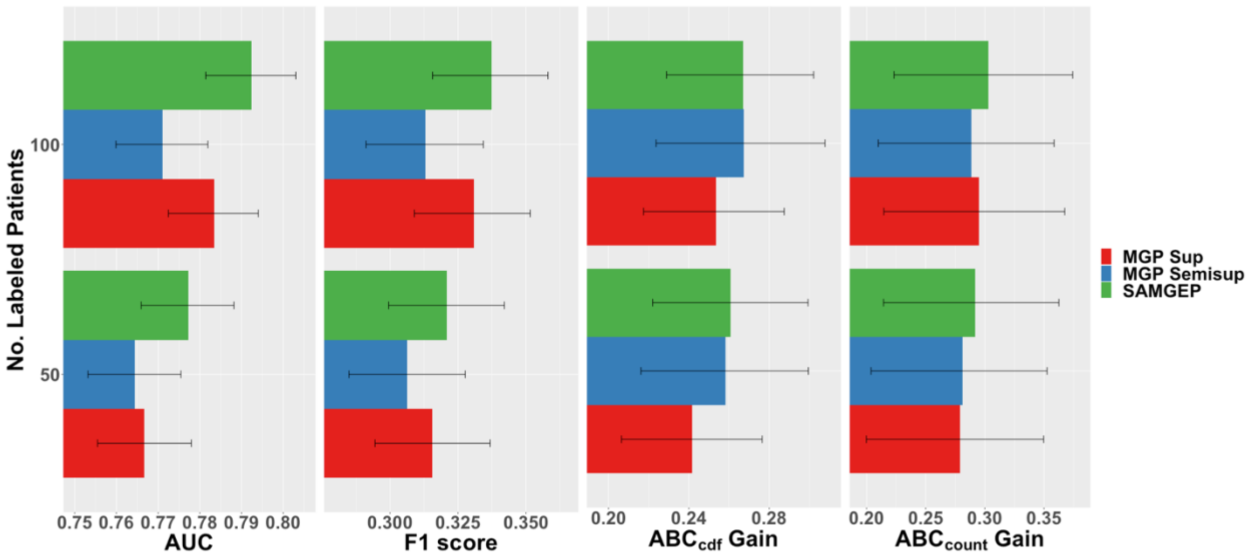


**Figure S1:** AUCs, F1 scores, $ABC_{cdf}$ gains, and $ABC_{count}$ gains for SAMGEP versus supervised and unsupervised MGP predicting MS relapse using real-world EHR data. 95% confidence intervals were empirically estimated by bootstrapping with 100 replicates.
